# Very low prevalence of validated *kelch13* mutations and absence of *hrp2/3* double gene deletions in South African malaria-eliminating districts (2022-2024)

**DOI:** 10.1101/2025.03.31.25324948

**Authors:** Jaishree Raman, Maxwell Mabona, Qedusizi Nyawo, Brighton Mangena, Gerdalize Kok, Lihle Mathaba, Gillian Malatje, Sonja B Lauterbach, Takalani I Makhanthisa, Hazel Gwarinda, Blaženka D Letinić, Arinao Ndadza, Eric Raswiswi, Mbavhalelo Shandukani, Ednah Baloyi, Ziyanda Fekema, Arshad Ismail, Jonathan Featherston, Bryan Greenhouse, Jennifer L Smith, Andres Aranda-Diaz

## Abstract

South Africa is one of 25 countries identified by the World Health Organization as having the potential to eliminate malaria in the near future. In response to the emerging threat of antimalarial resistance, South Africa enhanced its surveillance programs to enable the mapping of resistance markers prevalence down to the facility level. A total of 4 471 samples, collected between January 2022 and August 2024 from healthcare facilities, and during active surveillance in malaria-geliminating districts in KwaZulu-Natal and Mpumalanga provinces, were assessed for drug (mutations in the *kelch13*, *crt*, *mdr1*, *dhfr,* and *dhps* genes) and diagnostic (*hrp*2/3 gene deletions) resistance markers using PCR and sequencing (Sanger and/or targeted amplicon) protocols. Validated markers of artemisinin partial resistance were rare, with the P574L mutation detected as a minor allele in two samples and the P553L mutation present in one sample. Of the 60 additional non-synonymous mutations detected, the A578S (30/999) and the I494V (13/951) mutations were most prevalent. Almost all parasites assessed carried the *crtK76* (99.8%) and *mdr1N*86 (99.0%) alleles, suggesting susceptibility to chloroquine. The *dhfr* triple (99.9%) and *dhps* double (98.2%) mutations associated with pyrimethamine and sulfadoxine resistance, respectively, were close to fixation. No double *hrp*2/3 gene deletions were detected. These findings suggest that the recommended treatments and diagnostics in South Africa are effective. However, the strong selection for antimalarial drug resistance markers across southern Africa combined with high regional interconnectedness, emphasizes the need for sustained malaria molecular surveillance to support South and southern Africa achieve their elimination goals.

## Introduction

The World Health Organization (WHO) acknowledged the sustained progress made by South Africa in reducing its malaria burden through inclusion in its E2025 initiative.^1^ This initiative prioritized 25 malaria-endemic countries to receive additional support and technical guidance in their pursuit of halting indigenous malaria transmission within their national boundaries by 2025. However, several programmatic, biological, and environmental factors, including the recent emergence and spread of diagnostic^2^ and drug-resistant^3^ parasites have thwarted South Africa’s and other African countries’ ability to achieve this goal.

Artemisinin-based combination therapies (ACTs) are the WHO-recommended and preferred malaria treatment globally.^4^ Most infections in South Africa and Africa are treated with the ACT artemether-lumefantrine (Coartem®),^5^ which has been highly effective on the continent for over 20 years.^6^ African *Plasmodium falciparum* parasites with mutations in the propeller domain of the *kelch13* gene and confirmed delayed parasite clearance, the artemisinin partial resistance phenotype, are now entrenched in several countries from East, Central, and the Horn of Africa.^7^ Although delayed parasite clearance alone may have limited clinical relevance, due to the action of the partner drug, increased drug pressure heightens the risk of partner drug resistance emerging. Once partner drug resistance emerges, treatment failures are very likely, as seen in the Greater Mekong region.^8^

Given the devastating impact that ACT-resistant parasites would have on the malaria-vulnerable African population, the WHO released an Africa-specific response strategy.^6^ Routine surveillance for drug susceptibility using therapeutic efficacy studies and monitoring the prevalence of molecular markers of resistance are core activities of this strategy. The need for surveillance has become more urgent following the confirmation of parasites carrying *kelch13* mutations and deletions in the *histidine-rich protein* 2 and 3 (*hrp*2 and 3) coding genes in the Horn of Africa.^9^ These deletions enable parasites to evade detection by widely used HRP2-based rapid diagnostic tests (RDTs),^10-11^ preventing malaria diagnosis and treatment and facilitating the rapid spread of diagnostic and/or drug-resistant parasites.

South Africa established a genomic surveillance program^12^ following a drug-resistant malaria epidemic during the 1999/2000 malaria season.^13^ The findings directly led to artemether-lumefantrine replacing sulfadoxine-pyrimethamine (SP) as the first-line treatment.^14^ This surveillance program was further strengthened in 2018 by facilitating the linking of drug resistance profiles and individual patient data and the mapping of resistance data down to the healthcare facility level in malaria-eliminating districts in the provinces of KwaZulu-Natal and Mpumalanga (Figure 1) to support malaria-elimination activities.^15^ Surveillance for *hrp2/3* gene deletions was added to the surveillance program in 2022.

**Figure 1:**
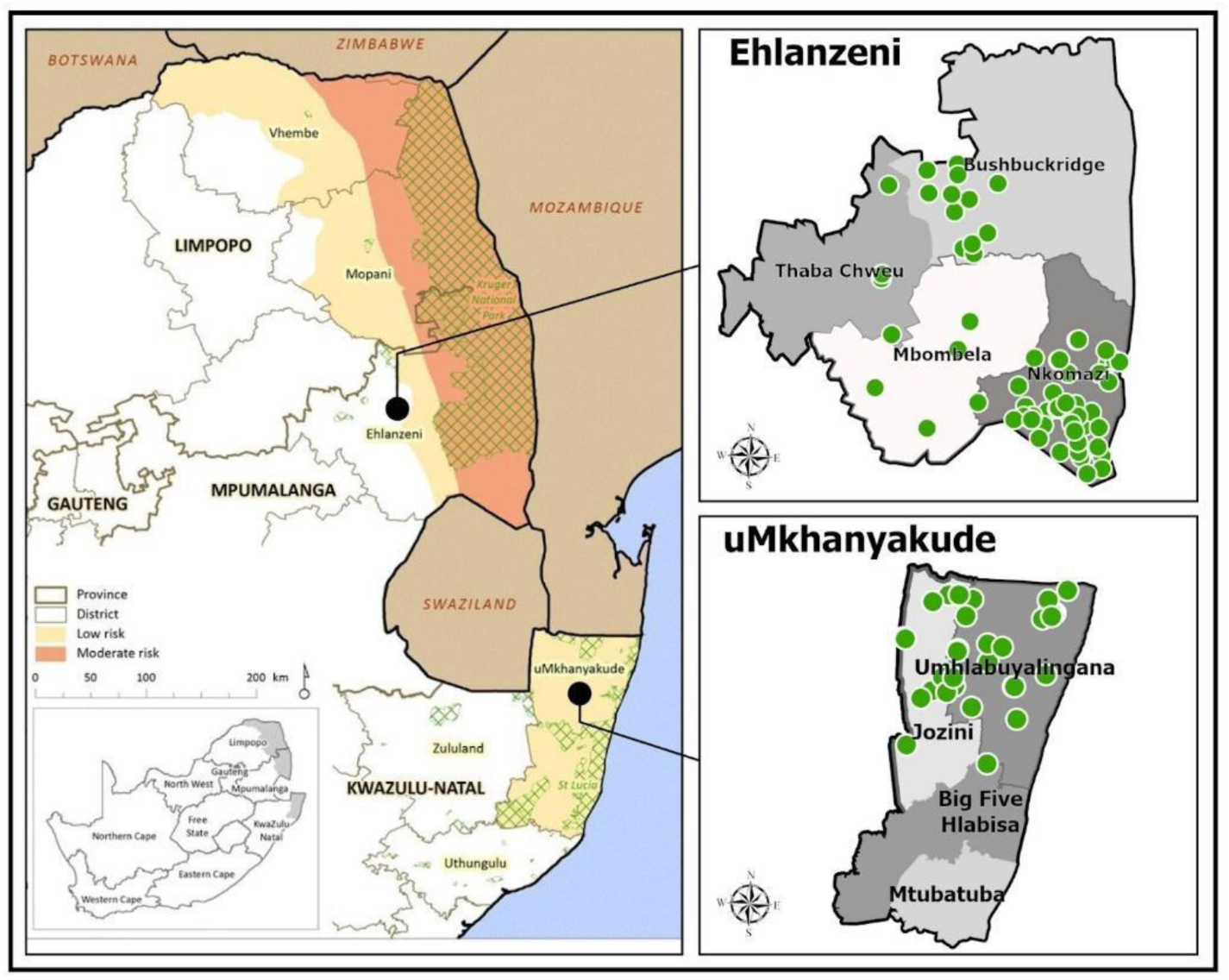
Malaria transmission areas in South Africa with the healthcare facilities (highlighted in green) sampled from a) Ehlanzeni and b) uMkhanyakude district municipalities.

This study presents findings on the prevalence of antimalarial drug and diagnostic resistance in malaria-eliminating districts of KwaZulu-Natal and Mpumalanga provinces during the 2022/2023 and 2023/2024 malaria seasons using published PCR and next-generation sequencing (NGS) protocols. The NGS data provided deeper insights into the genomic epidemiology of malaria in these eliminating districts. Findings from this study will be used to inform diagnostic and treatment policies in these districts, supporting malaria elimination efforts in South Africa.

## Methods and materials

### Country Setting

At the southernmost limit of malaria transmission in Africa, malaria in South Africa is restricted to the low-altitude tropical border eastern regions of three provinces Limpopo, Mpumalanga, and KwaZulu-Natal, bordering Botswana, Mozambique, and Zimbabwe (Figure 1). Sustained implementation of effective vector control (primarily indoor residual spraying) and case management interventions, supported by rigorous surveillance, have significantly reduced malaria transmission, allowing the country to transition to an elimination agenda in 2012.^13^ Approximately 14.4% of the population resides in malaria-risk areas,^16^ where transmission risk ranges from very low to moderate depending on locality.^17^ Most cases occur during the hot, humid wet months from September to March, with over 90% of confirmed cases due to *Plasmodium falciparum* infections.^18^ The dominant vector is *Anopheles arabiensis*,^19^ while the secondary vector, *An. merus* contributes to transmission in certain areas.^20^

All individuals in malaria-endemic areas with a fever or a recent history of fever are tested for malaria either by HRP2-based falciparum-specific RDTs at primary healthcare facilities and during active surveillance in the communities or by microscopy at the hospital level.^21^ Following confirmation of a *P. falciparum* infection, patients are treated with the ACT, artemether-lumefantrine for uncomplicated malaria, or intravenous artesunate for severe malaria.^18^ In selected eliminating areas, a single low-dose of the transmission-blocking antimalarial, primaquine, is co-administered with the first dose of artemether-lumefantrine to all eligible and consenting individuals.^18^

Malaria is a Category 1 notifiable condition in South Africa, requiring all cases to be reported within 24 hours of diagnosis through the Notifiable Medical Conditions (NMC) and District Health Information System 2 (DHIS2) platforms. In addition, basic patient demographic and travel information are captured on these platforms. In eliminating districts, cases should be investigated within 48 hours, with dried blood spots (DBS) collected from patients, where possible, on days 3 and 28 to monitor treatment efficacy.

### Study sites and sample collection

A total of 127 healthcare facilities, 32 and 95 from eliminating districts of uMkhanyakude district municipality, KwaZulu-Natal, and Ehlanzeni district municipality, Mpumalanga, respectively, participated in ongoing data collection for surveillance studies during the 2022/2023 and 2023/2024 seasons. Filter paper (Whatman Filter No 1 or TFN blood cards) finger-prick DBS from all RDT (First Response, Premier Technologies, India) malaria-positive individuals were collected by healthcare professionals based at healthcare facilities and malaria program staff during active case detection activities in the community. Routinely collected patient demographic and travel history data were linked to malaria-positive RDTs and associated DBS through a unique barcode. Samples (malaria-positive RDTs and associated DBS) were shipped weekly from Mpumalanga and monthly from KwaZulu-Natal to the central malaria molecular laboratory at the National Institute for Communicable Diseases (NICD) in Johannesburg, Gauteng, South Africa for downstream genomic analysis. For quality assurance and *hrp2/3* gene deletion assessments, 20% of randomly selected malaria RDT-negative samples were shipped to the NICD with the RDT-positive samples.

### Molecular analysis

Parasite DNA, extracted from the collected RDTs or DBS using either a modified Chelex method^22^ or the Qiagen DNA mini extraction kit (Qiagen, Germany), was subjected to multiplex *Plasmodium* species PCR^23^ assay to confirm a malaria infection and identify infecting species, and a subsequent *P*. *falciparum varATS* quantitative PCR (qPCR)^24^ assay to confirm and quantify *P*. *falciparum* parasitemia. All PCR and/or qPCR falciparum-confirmed samples were added to the drug resistance workflow.

Polymorphisms in the *P*. *falciparum chloroquine resistance transporter* (*crt)* 72-76 amino acid haplotype and the *multidrug resistance 1* (*mdr1)* 86 amino acid were assessed using previously described qPCR^25^ and restriction fragment length polymorphism (RFLP)^26^ assays, respectively. Polymorphisms in the propeller domain of the *kelch13* gene were assessed by nested PCR as described in Talundzic *et al*.^27^ followed by Sanger sequencing (Inqaba technologies, South Africa). The resulting *kelch13* sequences were aligned against a reference *P. falciparum kelch13* gene (XM_001350122.1) using a BLAST search and the ClustalW multiple-sequence tool to detect polymorphisms after codon 400 in the propeller domain of the *kelch13* gene, the genetic region containing the mutations associated with delayed parasite clearance.^28^ The *crt* 72-76 haplotype qPCR and *mdr1* 86 nested PCR assays were run in duplicate, with a third assay performed on any discordant results. Codons were classified as either pure wild-type, pure mutant, or mixed (both mutant and wild-type genotypes present in an individual sample).

A randomly selected subset of the samples from both transmission seasons were also genotyped using the Multiplex Amplicons for Drug, Diagnostic, Diversity, and Differentiation Haplotypes using High throughput Targeted Resequencing (MAD^4^HatTeR) amplicon deep sequencing workflow.^29^ Briefly, primer pools D1.1, R1.1 or R1.2, and R2.1 were used to amplify 241 targets across the parasite genome, and the resulting libraries were sequenced on an Illumina NextSeq 2000 instrument (Illumina, San Diego, USA) using 150 base pair paired-end reads. The MAD^4^HatTeR bioinformatic pipeline was used to infer alleles across all targets.^29^ Pool R1.2 is a subset of primers in R1.1, and we only report SNPs present in pool R1.2. In addition to the genes genotyped by traditional PCR assays indicated above, SNPs in the following genes were assessed: *dihydrofolate reductase* (*dhfr), dihydropterate synthase (dhps)*, *multidrug resistance 2 (mdr2)*, *coronin*, *apicoplast ribosomal protein S10* (*arps10)*, *putative phosphoinositide-binding* (*pib7)*, *ferredodin* (*fd)*, *exonuclease* (*exo)* and PF3D7_1322700. Target sequences in the *lactose dehydrogenase (ldh)* gene of the *P*. *vivax*, *P*. *ovale,* and *P*. *malariae* genomes were used to identify these species and differentiate between *P*. *ovale wallikeri* and *P*. *ovale curtisi*. Microhaplotypes from highly heterozygous targets were used to estimate the complexity of infection (COI). Variations to the original protocol and bioinformatic pipeline are described in Eloff *et al*.^30^

A random selection of RDT and PCR falciparum-positive samples and all RDT malaria-negative but PCR falciparum-positive samples with a parasite concentration of ≥ 50 parasite/µl of blood were assessed for *hrp2/3* gene deletions using previously published qPCR assays.^31-32^

### Data management

Malaria case data were extracted from the District Health Information System 2 (DHIS2) at regular intervals to cover January 2022 to August 2024. The extracted data contained routinely collected case information, including patient demographic characteristics, travel history, and case classification (imported, local, locally imported). After data cleaning and deduplication, individual patient demographic data and their associated antimalarial drug resistance profiles were linked through unique barcodes. Where possible, samples with missing, incomplete, or incorrectly captured barcodes were linked using a probabilistic fuzzy matching technique (R packages Fuzzyjoin v0.1.6; Stringdist v0.9.12). Data were categorized to distinguish the 2022–2023 season (January 2022 to April 2023) and the 2023–2024 season (May 2023 to August 2024).

### Statistical analysis and spatial mapping

R Studio (using R version 4.3.2) was used to clean, manage, and analyze linked genomic and patient demographic data. Basic descriptive statistical analysis was conducted using summary statistics and measures of central tendency. The Pearson χ2 test was used to test for statistically significant differences (p-value <0.05) between and within seasons and provinces.

ArcGIS Pro was used to map the two districts where samples were collected, including healthcare facility locations. Shapefiles were sourced from the open-source ArcGIS platform (pro.arcgis.com, 2023)

Drug resistance prevalence was estimated as the fraction of samples carrying a marker or mutation of the total number of positive samples with a valid genotype in the marker of interest. The prevalence of mixed genotypes was also calculated for every combination of genotypes observed in the same sample. To evaluate the association of patient demographic factors and genotypes of interest, a multivariate logistic regression analysis was conducted using a generalized linear model with a binomial distribution. The presence of a genotype was used as the binary outcome, and province, season, gender, age, and case classification were included as independent variables. This analysis was only performed for genotypes (*dhps*K540, *mdr1*Y184F, and *mdr2*I492V) and demographic categories (age, gender, and case classification) with sufficient samples for model convergence.

The complexity of infection (COI), and effective COI (eCOI, which accounts for within-sample relatedness) for each sample and population allele frequencies were estimated using the R package MOIRE (v3.4.0), which implements a Markov chain Monte Carlo-based approach to jointly infer sample COI and within host relatedness, and population allele frequencies, using polyallelic genomic data. By modeling the genetic data and accounting for experimental errors, MOIRE provides probabilistic estimates. Mean COI and the proportion of polyclonal infections within each population, along with their 95% Confidence Intervals (CI), were derived from the posterior distributions in MOIRE’s output. To assess associations of COI and eCOI with demographic groups, we performed multivariate analysis using a generalized linear model with a Poisson distribution. COI-1 or eCOI-1 was used as the outcome, and province/season, age group, parasitemia, sample sequencing depth, and case classification as independent variables. Differences in the percentage of polyclonal samples were also assessed with the same approach but with a binomial distribution with samples classified as polyclonal if they had an individual probability of polyclonality >0.05.

### Ethics approval

Approval for this surveillance study was obtained from the Human Research Ethics Committee: Medical of the University of Witwatersrand (M201124), the Mpumalanga Provincial Department of Health (MP_2015RP53_229), the KwaZulu-Natal Provincial Department of Health (KZ_202010_035), the South Africa National Department of Health and the University of California San Francisco Institutional Review Board (350074).

## Results

### Samples received

Over the study period, 49 122 RDT-positive and negative samples were received by the laboratory from 127 healthcare facilities, 23 415 and 25 707 during the 2022/2023 and 2023/2024 seasons, respectively (Figure S1). Most samples in each season (19 322 and 19 703) were submitted from the Ehlanzeni district municipality in Mpumalanga province. Of the 4 569 RDT-positive and 10 470 RDT-negative samples entered in the analytical workflow, 80.4 % and 6.0% were confirmed as falciparum-positive by qPCR, respectively (Table 1, Figure S1). During the 2023/2024 season, more RDT-negative samples were found to be malaria positive by PCR in both provinces (Table 1). Most (94.67%, 640/676) of the RDT-negative but PCR-positive samples had concentrations of <50 parasites/µl blood (Figure S2).

**Table 1:**
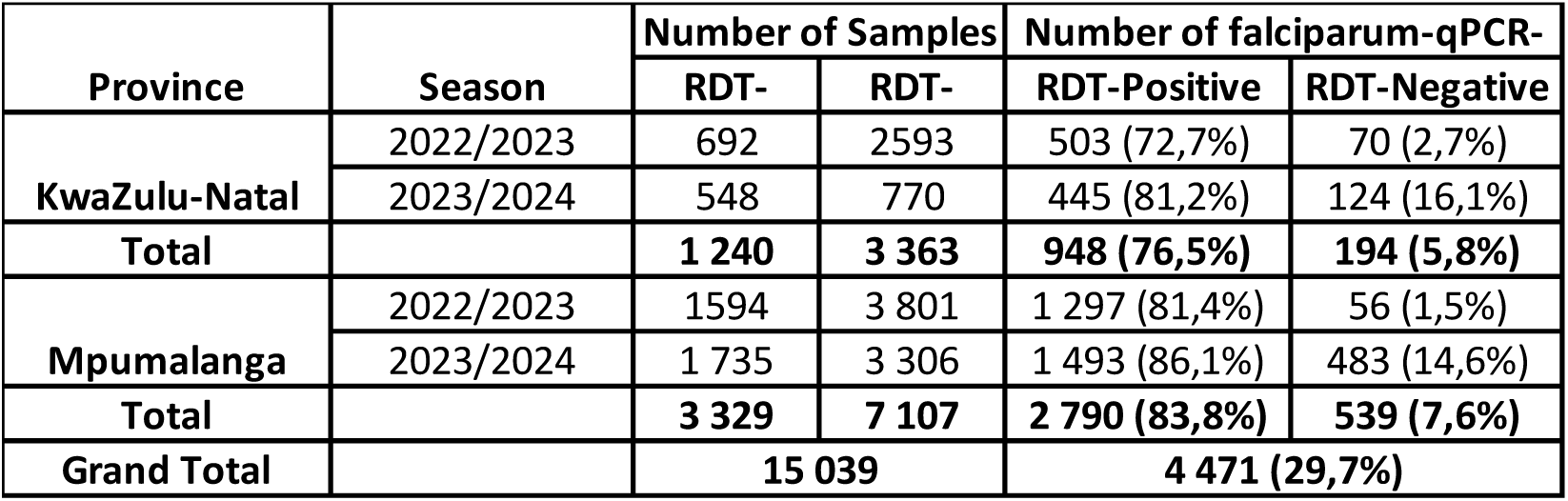
Number of rapid diagnostic tests/dried blood spots collected from KwaZulu-Natal and Mpumalanga provinces, South Africa, during the 2022/2023 and 2023/2024 malaria seasons and the number confirmed as falciparum-positive by qPCR.

Standard PCR assays detected only *P*. *falciparum* and *P*. *ovale* coinfections in less than 0.1% of the 4 471 *P*. *falciparum* qPCR positive samples. In contrast, 51 of 2 405 samples processed with the MAD^4^HatTeR protocol corresponded to *P*. *falciparum* coinfections with *P*. *malariae*, *P*. *ovale curtisi* and *P*. *ovale wallikeri* (Table S1). Most coinfections were detected in KwaZulu-Natal province, where they reached 9% of the samples, compared to 2% of samples from Mpumalanga province.

### Case characteristic

Except for the collection method, socio-demographic data for confirmed falciparum cases were often incomplete (Table 2). Most of the falciparum-confirmed samples received by the central laboratory over the two malaria seasons were from males (58.9%) aged 10 years and above (Table 2). The sample collection method was significantly different between the two provinces; 72% of the cases in Mpumalanga province were detected through passive surveillance, while 77% of the KwaZulu-Natal cases were collected through active case detection (p=<0.001, Table 1). Both provinces reported high numbers of imported cases, with imported cases slightly more common in KwaZulu-Natal (94%) compared to Mpumalanga (90%, p=0.014). Although citizenship was not frequently captured, when it was, most participants were Mozambican (70.1%), followed by South African (29.4%, Table 2). In KwaZulu-Natal province, the proportion of imported and actively detected cases significantly increased from 2022/23 to 2023/24, whereas Mpumalanga province showed the opposite trend.

**Table 2:** Socio-demographic characteristics of the confirmed falciparum-positive study population in KwaZulu-Natal and Mpumalanga provinces, South Africa, during the 2022/2023 and 2023/2024 malaria seasons.

| Socio-demographic characteristics | PROVINCE |  |  |  |  |  | OVERALL |  |  |
| --- | --- | --- | --- | --- | --- | --- | --- | --- | --- |
|  | KWAZULU-NATAL |  |  | MPUMALANGA |  |  |  |  |  |
|  | 2022/2023<br>(N=573) | 2023/2024<br>(N=569) | p-value | 2022/2023<br>(N=1,350) | 2023/2024<br>(N=1,979) | p-value | KZN<br>(N=1,142) | MPN<br>(N=3,329) | p-value |
| Gender |  |  |  |  |  |  |  |  |  |
| Female | 200 (47%) | 199 (38%) | 0.011 | 558 (44%) | 440 (37%) | <0.001 | 399 (42%) | 998 (41%) | 0.4 |
| Male | 229 (53%) | 319 (62%) |  | 715 (56%) | 741 (63%) |  | 548 (58%) | 1,456 (59%) |  |
| Unknown | 144 | 51 |  | 77 | 798 |  | 195 | 875 |  |
| Age catergy (Years) |  |  |  |  |  |  |  |  |  |
| 0 -<5 | 17 (3.1%) | 25 (6.0%) | 0.3 | 139 (11%) | 120 (7.7%) | <0.001 | 42 (4.4%) | 259 (9.3%) | <0.001 |
| 5 - <10 | 63 (12%) | 44 (11%) |  | 156 (13%) | 266 (17%) |  | 107 (11%) | 422 (15%) |  |
| 10 - <20 | 218 (40%) | 163 (39%) |  | 386 (32%) | 491 (31%) |  | 381 (40%) | 877 (31%) |  |
| 20 - <30 | 121 (22%) | 92 (22%) |  | 266 (22%) | 270 (17%) |  | 213 (22%) | 536 (19%) |  |
| 30 - <40 | 114 (21%) | 78 (19%) |  | 206 (17%) | 321 (21%) |  | 192 (20%) | 527 (19%) |  |
| ≥40 | 13 (2.4%) | 13 (3.1%) |  | 72 (5.9%) | 96 (6.1%) |  | 26 (2.7%) | 168 (6.0%) |  |
| Unknown | 27 | 154 |  | 125 | 415 |  | 181 | 540 |  |
| Case classification |  |  |  |  |  |  |  |  |  |
| Imported | 193 (89%) | 284 (97%) | <0.001 | 946 (93%) | 1,077 (87%) | <0.001 | 477 (94%) | 2,023 (90%) | 0.014 |
| Local | 24 (11%) | 8 (2.7%) |  | 68 (6.7%) | 152 (12%) |  | 32 (6.3%) | 220 (9.8%) |  |
| Locally imported | 0 (0%) | 0 (0%) |  | 1 (<0.1%) | 9 (0.7%) |  | 0 (0%) | 10 (0.4%) |  |
| Unknown | 356 | 277 |  | 335 | 741 |  | 633 | 1,076 |  |
| Method of detection |  |  |  |  |  |  |  |  |  |
| Active | 412 (72%) | 466 (82%) | <0.001 | 437 (32%) | 506 (26%) | <0.001 | 878 (77%) | 943 (28%) | <0.001 |
| Passive | 161 (28%) | 103 (18%) |  | 913 (68%) | 1,465 (74%) |  | 264 (23%) | 2,378 (72%) |  |
| Unknown |  |  |  | 0 | 8 |  | 0 | 8 |  |
| Citizenship |  |  |  |  |  |  |  |  |  |
| Mozambique | 167 (91%) | 48 (98%) | 0.4 | 10 (67%) | 10 (67%) | >0.9 | 215 (93%) | 20 (67%) | <0.001 |
| South Africa | 15 (8.2%) | 1 (2.0%) |  | 5 (33%) | 4 (27%) |  | 16 (6.9%) | 9 (30%) |  |
| Eswatini | 0 (0%) | 0 (0%) |  | 0 (0%) | 1 (6.7%) |  | 0 (0%) | 1 (3.3%) |  |
| Malawi | 1 (0.5%) | 0 (0%) |  | 0 (0%) | 0 (0%) |  | 1 (0.4%) | 0 (0%) |  |
| Unknown | 390 | 520 |  | 1,335 | 1,964 |  | 910 | 3,299 |  |

### Molecular analysis

Of the 4 471 qPCR falciparum-positive samples, 3 874 were assayed for *kelch13* polymorphisms by Sanger sequencing, with 2 700 (70%) yielding valid results; 2 178 were assayed for the *crt* 72-76 haplotype by qPCR, with 1 526 (70%) yielding valid results; and 1 054 were assayed for *mdr1* N86Y by RLFP, with 473 (45%) yielding a valid result. A total of 2 405 confirmed falciparum samples were added to the MAD^4^HatTeR workflow, of which 1 791 (74.5%) yielded a genotype for at least 15 SNPs of interest. The median per sample sequencing depth was 363 984 (IQR: 78 385-1 583 111), with a median per target depth of 646 (IQR 70-3 844). A median of 213 out of 238 (IQR 122-225) targets had more than 100 reads. The SNPs with the lowest genotyping rates were *crt* 218 and 220, with genotypes obtained from 549 samples; PF3D7_1322700 236, with 1 166 samples; and *mdr1* 86, with 1 220 samples. The demographic characteristics of the successfully sequenced samples resembled those of the falciparum-confirmed samples (Table S2).

### kelch13 mutations

*Kelch13* sequence data were generated from 2 453 (57%) samples with either Sanger sequencing or MAD^4^HatTeR. Samples carrying *kelch13* mutations were rare, with 63 (2.65%) with non-synonymous mutations in one of 12 amino acids detected over the study period (Table 3, Table S3-4). Two validated markers^33^ of artemisinin-partial resistance were observed in Mpumalanga province during the 2023/2024 season. The P574L mutation was a minor allele in two samples, and the P553L mutation was a unique allele in one sample. The most common *kelch13* mutations were the A578S and the I494V mutations (Table 3). The A578S mutation was observed in both provinces and seasons in less than 6% of the samples and with allele frequencies below 1.5% while the I494V mutation was detected in KwaZulu-Natal province in both seasons and Mpumalanga province in 2022/2023 in less than 4% of the samples and with allele frequencies below 1% (Table 3, S3). The prevalence of all detected *kelch13* mutations is presented in Table S4.

**Table 3:** Prevalence of mutations in the *kelch13, crt, mdr1, dhfr,* and *dhps* genes in KwaZulu-Natal and Mpumalanga provinces, South Africa, during the 2022/2023 and 2023/2024 malaria seasons.

| Gene | Codon | PROVINCE |  |  |  |  |  |  |  |
| --- | --- | --- | --- | --- | --- | --- | --- | --- | --- |
|  |  | KWAZULU-NATAL |  |  |  | MPUMALANGA |  |  |  |
|  |  | 2022-2023 |  | 2023-2024 |  | 2022-2023 |  | 2023-2024 |  |
|  |  | n/N | % | n/N | % | n/N | % | n/N | % |
| ***k13*** | Any non-synonymous mutation | 9/258 | 3,49 | 18/279 | 6,45 | 24/391 | 6,14 | 28/856 | 3,27 |
|  | V494I | 3/147 | 2.04 | 7/182 | 3.85 | 3/198 | 1.52 | 0/424 | 0.00 |
|  | P553L | 0/69 | 0.00 | 0/151 | 0.00 | 0/171 | 0.00 | 1/371 | 0.27 |
|  | P574L | 0/175 | 0.00 | 0/182 | 0.00 | 0/210 | 0.00 | 2/432 | 0.46 |
|  | A578S | 5/175 | 2.86 | 8/182 | 4.40 | 11/210 | 5.24 | 6/432 | 1.39 |
| ***crt*** | F48L | 0/184 | 0 | 2/278 | 0.72 | 0/383 | 0 | 5/854 | 0.59 |
|  | CVMNK 72-76 CVIET | 0/225 | 0 | 0/279 | 0 | 0/389 | 0 | 3/857 | 0.35 |
| ***mdr1*** | N86Y | 0/114 | 0 | 0/168 | 0 | 11/289 | 3,81 | 1/645 | 0.16 |
|  | N86F | 0/114 | 0 | 0/168 | 0 | 12/289 | 4,15 | 0/645 | 0 |
|  | Y184F | 92/145 | 63.45 | 148/230 | 64.35 | 252/359 | 70.19 | 560/824 | 67.96 |
| ***mdr2*** | I492V | 45/130 | 34.62 | 58/210 | 27.62 | 94/317 | 29.65 | 235/723 | 32.5 |
| ***dhfr*** | N51I | 144/144 | 100 | 243/243 | 100 | 331/334 | 99,1 | 780/781 | 99,87 |
|  | C59R | 144/144 | 100 | 243/243 | 100 | 334/334 | 100 | 779/781 | 99.74 |
|  | S108N | 159/159 | 100 | 263/263 | 100 | 370/370 | 100 | 839/839 | 100 |
| ***dhps*** | S436A | 5/165 | 3,03 | 1/263 | 0,38 | 2/380 | 0,52 | 8/847 | 0,94 |
|  | S436F | 3/165 | 1,82 | 1/263 | 0,38 | 1/380 | 0.26 | 1/847 | 0,12 |
|  | A437G | 164/165 | 99.39 | 258/263 | 98.1 | 372/380 | 97.89 | 830/847 | 97.99 |
|  | K540E | 137/140 | 97.86 | 236/239 | 98.74 | 330/337 | 97.92 | 756/774 | 97.67 |
|  | A581G | 0/232 | 0 | 1/279 | 0.36 | 1/379 | 0.26 | 5/847 | 0.59 |
|  | A613S | 1/227 | 0.44 | 0/263 | 0 | 0/358 | 0 | 2/791 | 0,25 |
|  | A613V | 0/227 | 0 | 0/263 | 0 | 0/358 | 0 | 2/791 | 0,25 |

Sanger sequencing and MAD^4^HatTeR produced comparable results for *kelch13* mutations. Among 1 257 samples with results from both methods, 96% of the results were fully concordant. MAD^4^HatTeR detected a mixed genotype in 49 samples (3.9%) with a unique genotype as per Sanger sequencing, most of which had a within-sample allele frequency (WSAF) for the discordant allele below 50% (Figure S3). Only one sample showed a unique allele that differed between the two methods.

### Other drug-resistance mutations

Mutations in the *dfhr* and *dhps* genes were only assessed through the MAD^4^HatTeR pipeline. The prevalence of mutations in the *dfhr* and *dhps* genes associated with resistance to pyrimethamine and sulfadoxine, respectively, was high in both provinces across both seasons (Table 3-4, S3). All samples from KwaZulu-Natal province while 99% of the samples from Mpumalanga province carried the *dhfr* triple haplotype (*dhfr*N51I+*dhfr*C59R+*dhfr*S108N) associated with pyrimethamine resistance.^34^ The *dhfrI*164L, associated with very high levels of pyrimethamine resistance,^35^ was not detected in any of the samples analyzed over the study period.

**Table 4:** Proportion (%) of *dhps* and *dhfr* mutations as haplotypes in KwaZulu-Natal and Mpumalanga provinces, South Africa, during the 2022/2023 and 2023/2024 malaria seasons.

| Gene | Mutation | PROVINCE |  |  |  |  |  |  |  |
| --- | --- | --- | --- | --- | --- | --- | --- | --- | --- |
|  |  | KWAZULU-NATAL |  |  |  | MPUMALANGA |  |  |  |
|  |  | 2022-2023 |  | 2023-2024 |  | 2022-2023 |  | 2023-2024 |  |
|  |  | N | % | N | % | N | % | N | % |
| *dhps+dhfr* | <5 mutations | 145 | 2.07 | 240 | 1.67 | 336 | 2.68 | 762 | 2.62 |
|  | quintuple |  | 91.03 |  | 94.58 |  | 92.26 |  | 89.76 |
|  | sextuple |  | 0 |  | 0 |  | 0.6 |  | 0.52 |
|  | undetermined |  | 6.9 |  | 3.75 |  | 4.46 |  | 7.09 |
| *dhfr* | <3 mutations | 158 | 0 | 260 | 0 | 360 | 1.11 | 826 | 0.36 |
|  | triple |  | 100 |  | 100 |  | 98.89 |  | 99.64 |
|  | undetermined |  |  |  |  |  |  |  |  |
| *dhps* | < 2 mutations | 145 | 2.07 | 240 | 1.25 | 339 | 0.88 | 765 | 0.26 |
|  | double |  | 91.03 |  | 94.58 |  | 92.92 |  | 90.46 |
|  | triple |  | 0 |  | 0 |  | 0.59 |  | 0.65 |
|  | undetermined |  | 6.9 |  | 3.75 |  | 4.42 |  | 6.67 |

The *dhps* double haplotype (*dhps*A437G+*dhps*K540E) associated with sulfadoxine resistance in southern Africa,^36^ was found in over 90% of the samples assessed in both provinces (Table 3-4, S3). Up to 7% of the samples had an undetermined haplotype due to mixed microhaplotypes in polyclonal infections. The number of samples with the quintuple mutation (*dhfr*N51I+*dhfr*C59R+*dhfr*S108N+*dhps*A437G+*dhps*K540E) linked with SP treatment failures in East and southern Africa^37^ mirrored the *dhps* number (Table 3-4, S3), given the fixation of the *dhfr* triple haplotype. Less than 1% of the samples carried mutations at codon *dhps*436 or *dhps*581 (Table 3-4, S3).

Due to low genetic diversity in *dhps* and *dhfr* haplotypes, analysis of the association of mutations and demographic factors was limited to the *dhps*K540 wildtype genotype. This genotype was more likely to be found in local cases than imported cases, with an odds ratio (OR) of 3.58 (95% confidence interval (CI): 2.04-6.30, Table S5). Conversely, participants aged 40 or older were less likely to carry the genotype than participants aged 4 years or younger, with an OR of 0.32 (CI: 0.12-0.89). No association with gender or other age categories was found.

The *crt*K76 and *mdr*1N86 alleles associated with full susceptibility to chloroquine^38^ but reduced susceptibility to lumefantrine^39-40^ remained close to fixation over the two seasons in both provinces (Table 3, S3-4). Concordance between the two workflows was very high for the *crt* CVMNK 72-76 CVIET, with only 2 of 573 (0.35%) samples genotyped by both methods producing discordant results. Both samples were classified as a mixed genotype as per MAD4HatTeR and a pure wildtype genotype as per qPCR. There was 100% concordance between 55 samples genotyped for *mdr1*N86Y by PCR and MAD4HatTeR. The *mdr1*Y184F mutation was found in 67.5% of the samples (Table 3) and was less likely to be found in locally transmitted infections, with an OR of 0.62 (CI 0.42-1.16, Table S6), with no significant associations with age or gender. Finally, the *mdr2*I492V mutation was found in 31.3% of the samples (Table 3) and was more likely to be carried by participants aged 5 to 9 years, with an OR of 2.19 (CI: 1.05-4.56, Table S5). No significant associations with other age groups, gender, or case classification were found.

### hrp2/3 deletions

No *hrp2* and *hrp3* gene deletion*s* were detected in the 603 RDT-negative but PCR-positive samples.

### Within host genetic diversity

Overall, the mean COI was higher in KwaZulu-Natal province (3.14, CI: 3.03-3.23) than in Mpumalanga province (2.83 CI: 2.78-2.88). The eCOI was similar between the two provinces (KwaZulu-Natal: 2.04, CI 2.02-2.05; Mpumalanga: 2.03, CI 2.02-2.04). The proportion of polyclonal infections also did not differ significantly (KwaZulu-Natal province: 71.4%, CI 69.3-73.2%; Mpumalanga province: 68.5%, CI = 67.4-70.0%). Furthermore, the COI and eCOI in KwaZulu-Natal province were significantly higher in the 2022-2023 season than in the 2023-2024 season (Figure 2, Table S6), although the proportion of polyclonal infections was not significantly different. All three metrics were significantly lower in locally acquired cases compared to imported cases (Figure 2, Table S6). Multivariate analysis adjusting for sequencing read depth, parasitemia and age confirmed these results (Table S7). The analysis also showed that samples from participants older than 40 had lower COI and eCOI compared to those from participants under 5 years of age (Table S7).

**Figure 2:**
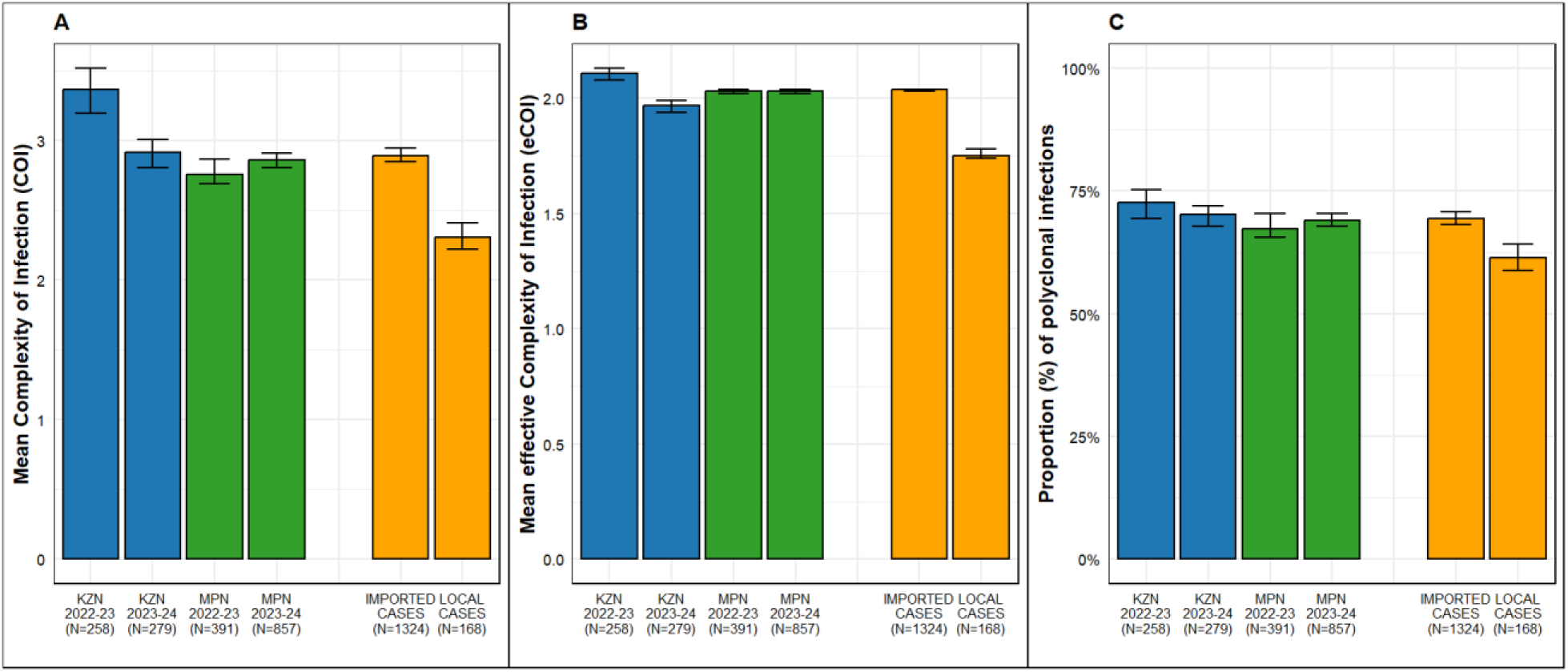
Complexity of infection (COI, A), effective COI (eCOI, B), and proportion of polyclonal infections in KwaZulu-Natal (KZN) and Mpumalanga (MPN) samples from the 2022/2023 and 2023/2024 malaria seasons, as well as in imported and local cases overall (C). Error bars represent 95% confidence intervals.

## Discussion

Despite over 15 years of sustained artemether-lumefantrine use in South Africa,^41^ the results from this study reveal limited selection for known markers of artemisinin-partial resistance. These findings confirm results from previous genomic surveillance^12^ and drug efficacy^42^ studies in the country. Mutations in the *kelch13* gene previously detected in South Africa^43-44^ have not remained in circulation, suggesting limited or no selective advantage. This differs from the trend observed in other southern African countries where *kelch13* mutations become established ^30,45-46^ and rapidly spread.^33,47^ However, the detection of two validated markers of artemisinin-partial resistance, *kelch13* P574L and P553L, in Mpumalanga during the 2023/2024 season is a concern. The prevalence of these and other novel mutations must be closely monitored and promptly responded to, should they become established and spread.

Polymorphisms in the *P. falciparum mdr1* gene modulate susceptibility to several antimalarials, including lumefantrine and amodiaquine.^48-49^ In areas with prolonged artemether-lumefantrine use, the *mdr1*N86 allele is selected for,^12,40^ a trend observed in this study. Parasites with this wildtype allele display lower ex vivo susceptibility to lumefantrine compared to parasites with mutant *mdr1*86Y allele.^50^ Although a meta-analysis of clinical trial data found an association between the *mdr1*N86 allele and treatment failures,^40^ the change in lumefantrine susceptibility linked to the *mdr1*N86 allele is modest,^51^ potentially requiring additional mutations for treatment failure.

While there have been no confirmed reports of treatment failure with artemether-lumefantrine in South Africa, data from two countries within the Southern African Development Community, Angola ^52^ and the Democratic Republic of Congo ^53^ reveal reduced artemether-lumefantrine efficacy. This is particularly concerning for South Africa. Like most African countries, artemether-lumefantrine is the first-line treatment for uncomplicated malaria,^6^ and currently, South Africa does not have a registered second-line ACT treatment available. South Africa must, therefore, intensify its efforts to protect the efficacy of artemether and lumefantrine. The safeguarding of these drugs has become more urgent as the next generation of antimalarials expected to come to market in the next two years contain lumefantrine (ganaplacide plus lumefantrine, artemether-lumefantrine plus amodiaquine).

As seen in other African countries, the withdrawal of chloroquine as the preferred treatment for uncomplicated falciparum malaria resulted in the selection of the wildtype *P. falciparum crt* K76 genotype in South Africa. This wildtype allele is associated with increased susceptibility to chloroquine.^54-55^ The re-emergence of chloroquine-susceptible parasites has ignited interest in re-introducing chloroquine for the treatment of falciparum malaria, particularly in areas where artemisinin-partial resistance has been confirmed. The prevalence of chloroquine-resistant parasites in and around the country should guide this decision, as chloroquine resistance will most likely be rapidly selected in areas where resistant parasites are present.

Even though SP has not been used in South Africa for malaria treatment or chemoprophylaxis since the late 2000s,^56^ mutations associated with resistance to sulfadoxine and pyrimethamine remain close to fixation.^12^ There are several potential drivers for the sustained selections of these mutations which include: 1) sustained regional SP drug pressure through its use in chemoprevention activities in many neighboring countries, 2) cross-reaction with cotrimoxazole, an antifolate-sulfonamide combination used to treat opportunistic infections, particularly in patients with HIV/AIDS^57^ and 3) the parasites circulating in South Africa carrying compensatory mutations which minimize the fitness costs of the SP mutations. The association of the wild type *dhps*K540 allele, linked to lower sulfadoxine resistance, with local transmission may reflect lower selective pressure in South Africa, compared to Mozambique, where most imported cases originate and SP chemoprevention is practiced. The reason for the higher prevalence of the mutant *dhps*540E allele in children under 5 is unclear. It may be a consequence of higher exposure to antifolate-sulfonamide-based drugs, but further research is required.

The WHO has reported that parasites carrying *hrp2* and *hpr3* gene deletions are rare in Africa, currently localized in the Horn of Africa.^11^ This study did not find any evidence of the selection for parasites carrying both the deletions, despite falciparum-specific RDTs being used at the primary healthcare level since 2001.^58^ While similar findings were reported from neighboring Mozambique,^59^ a recent study in KwaZulu-Natal with a small sample size ^60^ reported a high proportion of parasites with the *hrp2* and *hrp3* double deletion. Possible reasons for the discrepancy include the assessment of differing populations, sample quality, or differences in assay sensitivity and specificity. An in-depth investigation is urgently required to elucidate the cause of these differences. The large sample size in this study and the one in Mozambique^59^ support the continued use of HRP2-based-RDTs as the point-of-care diagnostic tool in South Africa. However, the prevalence of *hrp2/3* gene deletions must be closely monitored to ensure that the recommended point-of-care diagnostic can accurately detect malaria infections.

Parasite importation from higher-transmission areas can shape the genetic characteristics of local parasite populations. Higher transmission generally leads to greater population-level and intra-host genetic diversity. Consistent with this, imported infections in this study -accounting for 90% of cases, predominantly among males from Mozambique - had higher COI than locally transmitted infections. As a result, we found high levels of polyclonality in the eliminating districts of KwaZulu-Natal and Mpumalanga provinces, aligning with findings previously reported in neighboring Eswatini.^61^ Previous studies have shown that antimalarial drug-resistant parasites have been spread across Africa by highly mobile African populations.^37,62-63^ This highlights the increased risk of drug-resistant parasites being introduced in KwaZulu-Natal and Mpumalanga provinces through importation, as most cases notified in these provinces^64-65^ are imported. Although molecular markers of artemisinin-partial resistance are currently rare in South Africa, they are rapidly becoming established in several southern African countries including Namibia^30^and Zambia^45-46^ emphasizing the need for proactive routine genomic surveillance, particularly along known migration routes, to guide prompt mitigation responses.

As seen in earlier studies conducted in these two provinces,^64-65^ more males compared to females were affected by malaria, a trend observed in several other malaria-endemic countries.^66-67^ It is likely that work-related and social activities increase their exposure to malaria vectors. In addition, males are more likely to travel to malaria-endemic areas in search of work. This highlights the need for malaria awareness campaigns and interventions to target this high-risk group, particularly in malaria-eliminating areas.

Although all genotyping protocols produced comparable results, the MAD^4^HatTeR NGS workflow detected more minor alleles and non-falciparum co-infecting species, which could be attributed to differences in sensitivity. The clinical relevance of minor resistance alleles is currently unclear, so more research is required to understand their public health value. The high prevalence of falciparum co-infections with *P. ovale,* particularly in KwaZulu-Natal province, warrants further investigation. While artemether-lumefantrine can clear blood-stage ovale parasites, the antimalarial is ineffective against the dominant liver stage ovale parasite, resulting in relapsing malaria episodes. Further investigations are required to determine the true burden of non-falciparum *Plasmodium* in the country to guide case management responses.

This study has several limitations. First, resistance data were only generated from eliminating districts in Mpumalanga and KwaZulu-Natal provinces. No resistance data were generated for Limpopo Province, the province that reports the highest number of locally acquired cases.^68^ Drug- and diagnostic-resistant parasites may be present there, contributing to the sustained malaria burden in that province, underscoring the urgent need for implementing surveillance activities in Limpopo to ensure treatment effectiveness.

Second, only validated resistance markers were assessed, potentially missing novel mutations associated with reduced drug efficacy. A random selection of samples should be subjected to whole genome sequencing annually to proactively detect novel markers of resistance. Third, the absence of validated molecular markers for lumefantrine tolerance/resistance means currently, there is no molecular-based system for the early detection of reduced lumefantrine efficacy. Assessing parasite clearance on days 3 and 28 post-treatment is, therefore, essential for the prompt detection of potential treatment failures. Provincial programs must make every effort to collect these follow-up samples so that decreases in drug efficacy can be detected and responded to in a timely manner. Fourth, the poor quality of blood samples collected, particularly from KwaZulu-Natal province, limited the number of samples that could be analyzed using the NGS platform. Routine quality assessments of DBS collection should be conducted, with refresher training offered to facilities submitting suboptimal samples. Lastly, epidemiological analyses were hindered by missing critical data variables like travel history and sample barcodes in the malaria information systems. Healthcare workers and malaria program staff must be trained on the importance of complete and accurate patient information collection, with refresher training offered where needed.

## Conclusion

This study confirmed the limited selection of *kelch13* mutations associated with artemisinin-partial resistance and the absence of the *hrp2/3* double gene deletion associated with diagnostic resistance in eliminating districts of two South African malaria-endemic provinces. However, the continued selection for the reduced lumefantrine susceptibility marker, *mdr1*N86, in both provinces is concerning. Its entrenchment could potentially increase pressure on artemether to clear parasites, heightening the selection pressure for artemisinin-partial resistance in South Africa. This threat, together with the emergence and spread of *kelch13* mutations in several southern African countries, strengthens the call for routine malaria genomic surveillance in South Africa to guide timely responses to emerging resistance. The high prevalence of imported malaria in males needs to be addressed through targeted awareness campaigns and interventions.

## Data Availability

All data produced in the present study are available upon reasonable request to the authors

## Acknowledgments

We thank the study participants and communities for their willingness to participate in the study, Ongeziwe Taku and Nokwethemba Kubheka for assistance with sample processing, the healthcare professionals, staff from the Mpumalanga and KwaZulu-Natal Malaria Programmes, and the funders, without whose support this genomic surveillance study would not have been possible.

## Author’s Current Address

Jaishree Raman (JR): ARMMOR, National Institute for Communicable Disease, 1 Modderfontien Road, Sandringham, Gauteng, 2131, South Africa.

Maxwell Mabona (MM): ARMMOR, National Institute for Communicable Disease, 1 Modderfontien Road, Sandringham, Gauteng, 2131, South Africa.

Qedusizi Nyawo (QN): KwaZulu-Natal Provincial Malaria Programme, Nsinde Street, Jozini, KwaZulu-Natal, 3969, South Africa.

Brighton Mangena (BM): SADC Malaria Elimination Eight Secretariat, Windhoek, Namibia

Gerdalize Kok (GK): Mpumalanga Provincial Malaria Control Programme, 666 Anderson Street, Mbombela, Mpumalanga, 1200, South Africa.

Lihle Mathaba (LM): Mpumalanga Provincial Malaria Control Programme, 666 Anderson Street, Mbombela, Mpumalanga, 1200, South Africa.

Gillian Malatje (GM): Mpumalanga Provincial Malaria Control Programme, 666 Anderson Street, Mbombela, Mpumalanga, 1200, South Africa.

Sonja B Lauterbach (SL): ARMMOR, National Institute for Communicable Disease, 1 Modderfontien Road, Sandringham, Gauteng, 2131, South Africa.

Takalani I Makhanthisa (TIM): ARMMOR, National Institute for Communicable Disease, 1 Modderfontien Road, Sandringham, Gauteng, 2131, South Africa.

Hazel Gwarinda (HG): ARMMOR, National Institute for Communicable Disease, 1 Modderfontien Road, Sandringham, Gauteng, 2131, South Africa.

Blaženka D Letinić (BDL): Centre for Vaccines and Immunology, National Institute for Communicable Diseases, 1 Modderfontien Road, Sandringham, Johannesburg, Gauteng, 2131, South Africa.

Arinao Ndadza (AN): ARMMOR, National Institute for Communicable Disease, 1 Modderfontien Road, Sandringham, Gauteng, 2131, South Africa.

Eric Raswiswi (ER): Natalia Building, 330 Langalibalele Street, Pietermaritzburg, KwaZulu-Natal, 3201, South Africa.

Mbavhalelo B Shandukani (MBS):National Department of Health, 1112 Voortrekker Road, Pretoria Townlands, Gauteng, 0187, South Africa.

Ednah Baloyi (EB): National Department of Health, 1112 Voortrekker Road, Pretoria Townlands, Gauteng, 0187, South Africa.

Ziyanda Fekema (ZF): National Department of Health, 1112 Voortrekker Road, Pretoria Townlands, Gauteng, 0187, South Africa.

Arshad Ismail: Sequencing Core Facility, National Institute for Communicable Disease, 1 Modderfontien Road, Sandringham, Gauteng, 2131, South Africa.

Jonathan Featherston (JF): Sequencing Core Facility, National Institute for Communicable Disease, 1 Modderfontien Road, Sandringham, Gauteng, 2131, South Africa.

Bryan Greenhouse (BG): EPPIcenter, University of San Francisco California, 1001 Potrero Avenue, San Francisco, California, 94110, USA

Jennifer L Smith (JLS): Department of Epidemiology and Biostatistics, University of San Francisco California, 550 16th Street, San Francisco, California, 94158, USA

Andres Aranda-Diaz (AAD): ISGlobal, Carrer Rossello, 132 6-2, Barcelona, 08036, Spain.

## Author contributions

JR, BG, JLS: Designed the study

JR, HG, SBL, TIM, BDL, AN, AI, JF: Conducted the laboratory analysis and some data analysis

AAD, MM: Conducted the epidemiological and statistical analyses

MM, BM, GK, ZF: Managed the epidemiological databases

QN, LM, GM, ER, MBS, EB: Oversaw the management of sample collection and their shipment to the central laboratory

AAD: Developed bioinformatics pipelines and processed raw next-generation sequence data

JR, AAD: Analysed and interpreted the genomic data

JR: Drafted the manuscript with all co-authors providing input to the initial draft

All authors approved the submission of the final version.

## Supplementary Tables

**Table S1:**
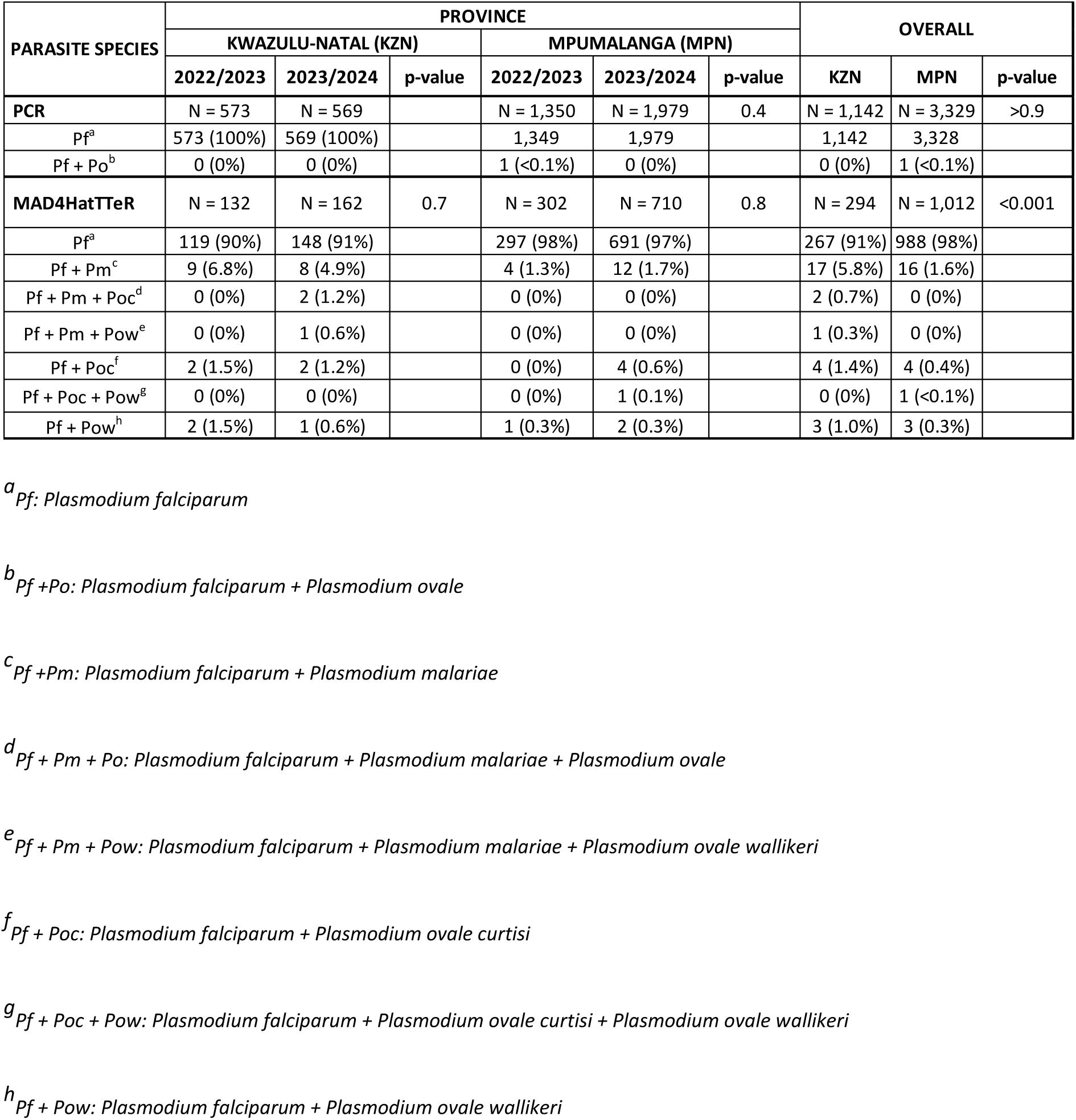
Malaria species identified by standard PCR and the MAD^4^HatTeR protocols in KwaZulu-Natal and Mpumalanga provinces, during the 2022/2023 and 2023/2024 malaria seasons.

**Table S2:** Cross-tabulation of socio-demographic characteristics between confirmed falciparum samples and samples successfully sequenced using MAD^4^HatTeR.

| Socio-demographic | Total qPCR<br>N = 2,338 (%) | Successfully<br>N = 1,791 (%) | p-value |
| --- | --- | --- | --- |
| **Province** |  |  |  |
| KwaZulu-Natal | 701 (30%) | 537 (30%) | >0.9 |
| Mpumalanga | 1,630 (70%) | 1,248 (70%) |  |
| Unknown | 7 | 6 |  |
| **Malaria Season** |  |  |  |
| 2022-2023 | 916 (39%) | 649 (36%) | <0.001 |
| 2023-2024 | 1,415 (61%) | 1,136 (64%) |  |
| Unknown | 7 | 6 |  |
| **Gender** |  |  |  |
| Female | 640 (40%) | 512 (41%) | 0.017 |
| Male | 975 (60%) | 730 (59%) |  |
| Unknown | 723 | 549 |  |
| **Age group** |  |  |  |
| 0 - <5 | 202 (8.8%) | 163 (9.3%) | 0.4 |
| 5 - <10 | 113 (4.9%) | 92 (5.2%) |  |
| 10 - <20 | 294 (13%) | 225 (13%) |  |
| 20 - <30 | 698 (30%) | 521 (30%) |  |
| 30 - <40 | 484 (21%) | 364 (21%) |  |
| ≥40 | 508 (22%) | 392 (22%) |  |
| Unknown | 39 | 34 |  |
| **Case classification** |  |  |  |
| Imported | 1,678 (88%) | 1,325 (88%) | 0.8 |
| Local | 210 (11%) | 170 (11%) |  |
| Locally imported | 13 (0.7%) | 11 (0.7%) |  |
| Unknown | 437 | 285 |  |

**Table S3:** Allele frequency for all genotypes assessed in falciparum samples from KwaZulu-Natal and Mpumalanga provinces, South Africa, during the <u>2022/</u>2023 and 2023/2024 malaria seasons.

| Gene | Amino acid | Allele | PROVINCE |  |  |  |
| --- | --- | --- | --- | --- | --- | --- |
|  |  |  | KWAZULU-NATAL |  | MPUMALANGA |  |
|  |  |  | 2022-2023 | 2023-2024 | 2022-2023 | 2023-2024 |
| *dhfr* | 16/51/59 | AIC | 0.000237 (3.47e-08 - 0.00249) | 0.00182 (4.76e-08 - 0.00694) | 0.00412 (0.000877 - 0.0103) | 0.00478 (0.00205 - 0.00873) |
| *dhfr* | 16/51/59 | AIR | 0.997 (0.989 - 1) | 0.998 (0.993 - 1) | 0.989 (0.981 - 0.995) | 0.988 (0.982 - 0.993) |
| *dhfr* | 16/51/59 | ANR | 0.00289 (4.79e-05 - 0.011) | 0 | 0.00666 (0.00232 - 0.0135) | 0.00707 (0.00376 - 0.0114) |
| *dhps* | 431/436/437 | IAA | 0.00712 (0.000772 - 0.0199) | 0.000107 (1.58e-12 - 0.00119) | 0.00129 (4.44e-05 - 0.00465) | 0.00349 (0.00127 - 0.00703) |
| *dhps* | 431/436/437 | IAG | 1.18e-11 (1.03e-12 - 7.13e-11) | 0 | 0 | 4.23e-05 (2.94e-08 - 0.000407) |
| *dhps* | 431/436/437 | IFA | 0.00266 (6.51e-05 - 0.0102) | 0.000134 (3.05e-08 - 0.00127) | 6.27e-05 (1.4e-12 - 0.000711) | 0 |
| *dhps* | 431/436/437 | IFG | 5.31e-12 (1.02e-12 - 3.23e-11) | 0 | 0 | 6.04e-05 (3.46e-08 - 0.000557) |
| *dhps* | 431/436/437 | ISA | 0.0496 (0.0315 - 0.0751) | 0.0215 (0.0107 - 0.0347) | 0.0316 (0.0207 - 0.0448) | 0.0463 (0.0357 - 0.0563) |
| *dhps* | 431/436/437 | ISG | 0.941 (0.914 - 0.961) | 0.978 (0.965 - 0.989) | 0.967 (0.953 - 0.978) | 0.95 (0.939 - 0.961) |
| *dhps* | 431/436/437 | ICA | 0 | 0.000144 (3.36e-08 - 0.00145) | 0 | 0 |
| *dhps* | 540/581 | EA | 0.946 (0.917 - 0.969) | 0.967 (0.948 - 0.981) | 0.952 (0.934 - 0.967) | 0.942 (0.93 - 0.953) |
| *dhps* | 540/581 | KA | 0.0543 (0.0313 - 0.0829) | 0.0331 (0.0186 - 0.0522) | 0.0468 (0.0323 - 0.0632) | 0.0558 (0.0454 - 0.068) |
| *dhps* | 540/581 | EG | 0 | 0 | 0.0014 (3.63e-05 - 0.00505) | 0.00182 (0.000359 - 0.00408) |
| *dhps* | 540/581 | ET | 0 | 0 | 0 | 7.84e-05 (3.25e-08 - 0.000703) |
| *dhps* | 613 | A | 1 ( 1 - 1) | 0 | 0 | 0.999 (0.997 - 1) |
| *dhps* | 613 | S | 4.69e-08 (3.32e-08 - 5.52e-08) | 0 | 0 | 0.000659 (2.01e-05 - 0.00245) |
| *dhps* | 613 | V | 0 | 0 | 0 | 0.00059 (1.84e-05 - 0.00242) |
| *k13* | 494 | I | 0.00224 (4.73e-05 - 0.00762) | 0.00935 (0.00284 - 0.0188) | 0.00347 (0.000554 - 0.00901) | 0.00115 (0.000159 - 0.00325) |
| *k13* | 494 | V | 0.998 (0.992 - 1) | 0.991 (0.981 - 0.997) | 0.997 (0.991 - 0.999) | 0.999 (0.997 - 1) |
| *k13* | 578 | A | 0.992 (0.983 - 0.998) | 0.986 (0.976 - 0.994) | 0.985 (0.975 - 0.993) | 0.995 (0.991 - 0.997) |
| *k13* | 578 | S | 0.00756 (0.00218 - 0.0168) | 0.0137 (0.00576 - 0.0245) | 0.0144 (0.00696 - 0.0244) | 0.00534 (0.00257 - 0.00929) |
| *k13* | 578 | T | 7.37e-05 (1.04e-12 - 0.000923) | 0 | 0.000128 (3.3e-08 - 0.00113) | 6.11e-05 (3.22e-08 - 0.000534) |
| *mdr1* | 182/184/186 | GFW | 0.432 (0.365 - 0.497) | 0.479 (0.421 - 0.532) | 0.494 (0.451 - 0.537) | 0.48 (0.449 - 0.51) |
| *mdr1* | 182/184/186 | GYW | 0.568 (0.503 - 0.635) | 0.521 (0.468 - 0.579) | 0.506 (0.463 - 0.549) | 0.52 (0.49 - 0.551) |
| *mdr1* | 86 | F | 0 | 0 | 8.08e-12 (1.02e-12 - 2.73e-11) | 0 |
| *mdr1* | 86 | N | 0 | 0 | 0.995 (0.988 - 0.999) | 1 ( 1 - 1) |
| *mdr1* | 86 | Y | 0 | 0 | 0.00516 (0.00115 - 0.0122) | 1.61e-09 ( 0 - 2.9e-08) |
| *mdr2* | 492 | I | 0.812 (0.762 - 0.862) | 0.832 (0.791 - 0.871) | 0.832 ( 0.8 - 0.862) | 0.811 (0.787 - 0.833) |
| *mdr2* | 492 | V | 0.188 (0.138 - 0.238) | 0.168 (0.129 - 0.209) | 0.168 (0.138 - 0.2) | 0.189 (0.167 - 0.213) |

**Table S4:** Prevalence of all *kelch13* mutation detection in KwaZulu-Natal and Mpumalanga provinces, South Africa, during the 2022/2023 and 2023/2024 malaria seasons.

| *kelch13*<br>mutation | PROVINCE |  |  |  |  |  |  |  |
| --- | --- | --- | --- | --- | --- | --- | --- | --- |
|  | KWAZULU-NATAL |  |  |  | MPUMALANGA |  |  |  |
|  | 2022/2023 |  | 2023/2024 |  | 2022/2023 |  | 2023/2024 |  |
|  | n/N | % | n/N | % | n/N | % | n/N | % |
| V494I | 3/147 | 2.04 | 7/182 | 3.85 | 3/198 | 1.52 | 0/424 | 0.00 |
| P553L | 0/69 | 0.00 | 0/151 | 0.00 | 0/171 | 0.00 | 1/371 | 0.27 |
| P574L | 0/175 | 0.00 | 0/182 | 0.00 | 0/210 | 0.00 | 2/432 | 0.46 |
| A578S | 5/175 | 2.86 | 8/182 | 4.40 | 11/210 | 5.24 | 6/432 | 1.39 |
| A578T | 1/175 | 0.57 | 0/182 | 0.00 | 0/210 | 0.00 | 1/432 | 0.23 |
| C580R | 0/175 | 0.00 | 1/182 | 0.55 | 1/210 | 0.48 | 0/432 | 0.00 |
| V581A | 0/175 | 0.00 | 2/182 | 1.10 | 0/210 | 0.00 | 0/432 | 0.00 |
| L598S | 0/174 | 0.00 | 1/178 | 0.56 | 1/203 | 0.49 | 0/416 | 0.00 |
| N599D | 0/174 | 0.00 | 0/178 | 0.00 | 0/203 | 0.00 | 1/416 | 0.24 |
| E605V | 0/174 | 0.00 | 1/178 | 0.56 | 0/203 | 0.00 | 0/416 | 0.00 |
| K607E | 0/174 | 0.00 | 1/178 | 0.56 | 1/203 | 0.49 | 0/416 | 0.00 |
| K607R | 0/174 | 0.00 | 0/178 | 0.00 | 3/203 | 1.48 | 2/416 | 0.48 |

**Table S5:** Multivariate logistic regression results for relevant demographics and drug resistance markers of interest.

| Characteristic | *dhps* K540 |  |  | *mdr1* Y184F |  |  | *mdr2* I492V |  |  |
| --- | --- | --- | --- | --- | --- | --- | --- | --- | --- |
|  | n/N (%) | OR<sup>a</sup> (95% CI<sup>b</sup>) | p-value | n/N (%) | OR<sup>a</sup> (95% CI<sup>b</sup>) | p-value | n/N (%) | OR<sup>a</sup> (95% CI<sup>b</sup>) | p-value |
| **Gender** | **N=950** |  |  | **N=1004** |  |  | **N=885** |  |  |
| Female | 35/396 (8.83%) | Ref<sup>c</sup> |  | 281/416 (67.54%) | Ref<sup>c</sup> |  | 104/361 (28.80%) | Ref<sup>c</sup> |  |
| Male | 51/554 (9.20%) | 1.09 (0.68 - 1.73) | 0,7 | 406/588 (69.04%) | 1.06 (0.80 - 1.39) | 0,7 | 169/524 (32.25%) | 1.19 (0.88 - 1.60) | 0,3 |
| **Age Group** |  |  |  |  |  |  |  |  |  |
| 0-4 | 10/105 (9.52%) | Ref<sup>c</sup> |  | 79/111 (71.17%) | Ref<sup>c</sup> |  | 23/96 (23.96%) | Ref<sup>c</sup> |  |
| 5-9 | 6/57 (10.52%) | 0.68 (0.22-2.09) | 0,5 | 49/64 (76.56%) | 1.50 (0.73 - 3.09) | 0,3 | 22/53 (41.51%) | 2.19 (1.05 - 4.56) | **0,037** |
| 10-19 | 22/140 (15.71%) | 1.34 (0.59 - 3.04) | 0,5 | 97/140 (69.28%) | 1.01 (0.58 - 1.76) | >0.9 | 42/125 (33.60%) | 1.58 (0.86 - 2.90) | 0,14 |
| 20-29 | 22/278 (7.91%) | 0.82 (0.37 - 1.82) | 0,6 | 206/291 (70.79%) | 0.98 (0.60 - 1.59) | >0.9 | 75/266 (28.20%) | 1.20 (0.70 - 2.06) | 0,5 |
| 30-39 | 19/191 (9.94%) | 0.97 (0.43 - 2.21) | >0.9 | 130/197 (65.98%) | 0.81 (0.49 - 1.35) | 0,4 | 61/176 (34.66%) | 1.69 (0.96 - 2.97) | 0,069 |
| 40+ | 7/179 (3.91%) | 0.32 (0.12 - 0.89) | **0,029** | 126/201 (62.68%) | 0.70 (0.42 - 1.16) | 0,2 | 50/169 (29.59%) | 1.33 (0.75 - 2.37) | 0,3 |
| **Case classification** |  |  |  |  |  |  |  |  |  |
| Imported | 61/834 (7.31%) | Ref<sup>c</sup> |  | 616/883 (69.76%) | Ref<sup>c</sup> |  | 238/779 (30.55%) | Ref<sup>c</sup> |  |
| Local | 24/110 (21.81%) | 3.58 (2.04 - 6.30) | **<0.001** | 68/115 (59.13%) | 0.62 (0.40 - 0.94) | **0,023** | 35/100 (35.00%) | 1.06 (0.67 - 1.69) | 0,8 |
| Locally imported | 1/6 (16.66%) | 3.54 (0.38 - 32.6) | 0,3 | 3/6 (50.00%) | 0.50 (0.10 - 2.53) | 0,4 | 0/6 (0.00%) | 0.00 (0.00 - inf) | >0.9 |

<sup>a</sup>OR: Odds Ratio<sup>b</sup>95% CI: 95% Confidence Interval<sup>c</sup>Ref: Reference group

**Table S6:** Within host genetic diversity in samples in KwaZulu-Natal and Mpumalanga provinces, South Africa, during the 2022/2023 and 2023/2024 malaria seasons.

| Variable | PROVINCE |  |  |  | CASE CLASSIFICATION |  |
| --- | --- | --- | --- | --- | --- | --- |
|  | KWAZULU-NATAL |  | MPUMALANGA |  | Imported | LOCAL |
|  | 2022/2023 | 2023/2024 | 2022/2023 | 2023/2024 | N= 1322 | N = 170 |
| **COI<sup>a</sup>** | 3.37 (3.20-3.52) | 2.92 (2.81-3.01) | 2.76 (2.69-2.87) | 2.86 (2.81-2.91) | 2.89 (2.85-2.95) | 2.31 (2.22-2.41) |
| **eCOI<sup>b</sup>** | 2.11 (2.08-2.13) | 1.97 (1.94-1.99) | 2.03 (2.02-2.04) | 2.03 (2.02-2.04) | 2.04 (2.03-2.04) | 1.75 (1.74-1.78) |
| **% polyclonal** | 72.59% (69.38-75.19%) | 70.22% (67.74-72.05%) | 67.25% (65.47-70.33%) | 69.05% (67.79-70.48%) | 69.31 % (68.23-70.73%) | 61.42% (58.82-64.12%) |

<sup>a</sup>COI: complexity of infection
<sup>b</sup>eCOI: effective COI
Mean (95% confidence interval)

**Table S7:** Multivariate logistic regression results for relevant demographics and COI, eCOI and polyclonality.

| Outcome | COI<sup>a</sup> |  | eCOI<sup>b</sup> |  | Probability polyclonal >0.05 |  |
| --- | --- | --- | --- | --- | --- | --- |
| Coefficients | Estimate | p value | Estimate | p value | Estimate | p value |
| Intercept | 1,51E+00 | <0.001 | 8,49E-01 | <0.001 | 1,49E+00 | <0.001 |
| KZN-2023/2024 | -5,80E-01 | <0.001 | -4,73E-01 | 0,007 | -7,11E-01 | 0,083 |
| MPN-2022/2023 | -4,10E-01 | 0,001 | -2,13E-01 | 0,197 | -1,96E-01 | 0,623 |
| MPN-2023/2024 | -2,80E-01 | 0,010 | -2,14E-01 | 0,151 | -2,85E-01 | 0,446 |
| log10(parasitemia) | -1,77E-01 | <0.001 | -1,39E-01 | <0.001 | -1,03E-07 | 0,409 |
| Sequencing_depth | 2,71E-08 | 0,100 | 4,19E-08 | 0,035 | 2,34E-08 | 0,558 |
| Age:5-9 | 1,92E-01 | 0,186 | 1,49E-01 | 0,402 | NA | NA |
| Age:10-19 | -2,13E-02 | 0,869 | -1,33E-01 | 0,405 | NA | NA |
| Age:20-29 | -4,25E-03 | 0,970 | -1,23E-01 | 0,379 | NA | NA |
| Age:30-39 | -7,49E-02 | 0,530 | -2,47E-01 | 0,096 | NA | NA |
| Age:40+ | -2,43E-01 | 0,045 | -4,46E-01 | 0,004 | NA | NA |
| Local | -3,45E-01 | <0.001 | -2,80E-01 | 0,028 | -4,89E-01 | 0,033 |
| Locally imported | -1,30E-01 | 0,671 | -3,32E-01 | 0,459 | 3,48E-02 | 0,966 |

<sup>a</sup>COI: complexity of infection
<sup>b</sup>eCOI: effective COI.

## Supplementary Figures

**Figure S1:**
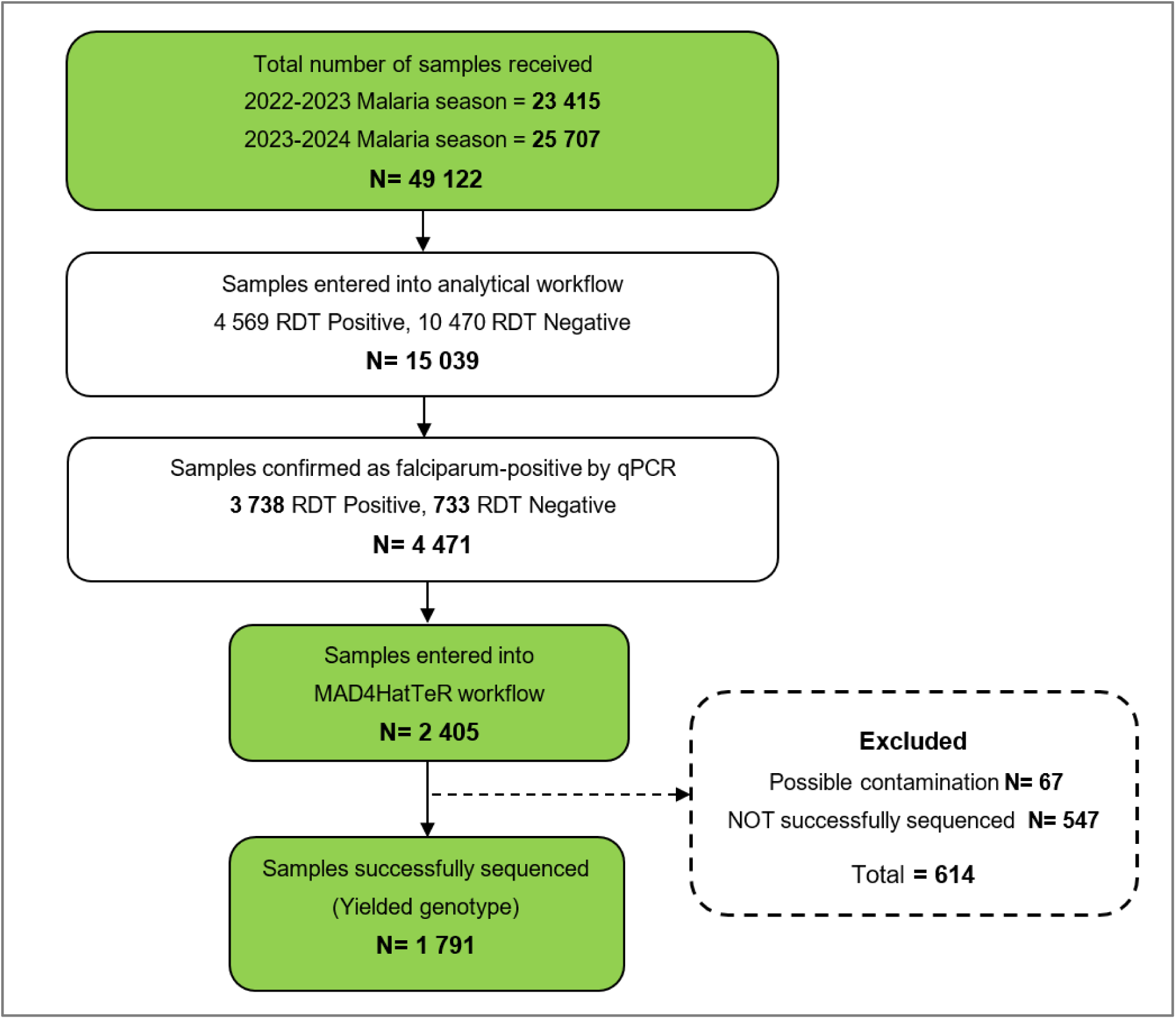
The number of samples received by the Central Laboratory during the 2022/2023 and 2023/2024 malaria seasons, the number entered into analytical workflow, and the number successfully sequenced through the MAD^4^HatTer workflow.

**Figure S2:**
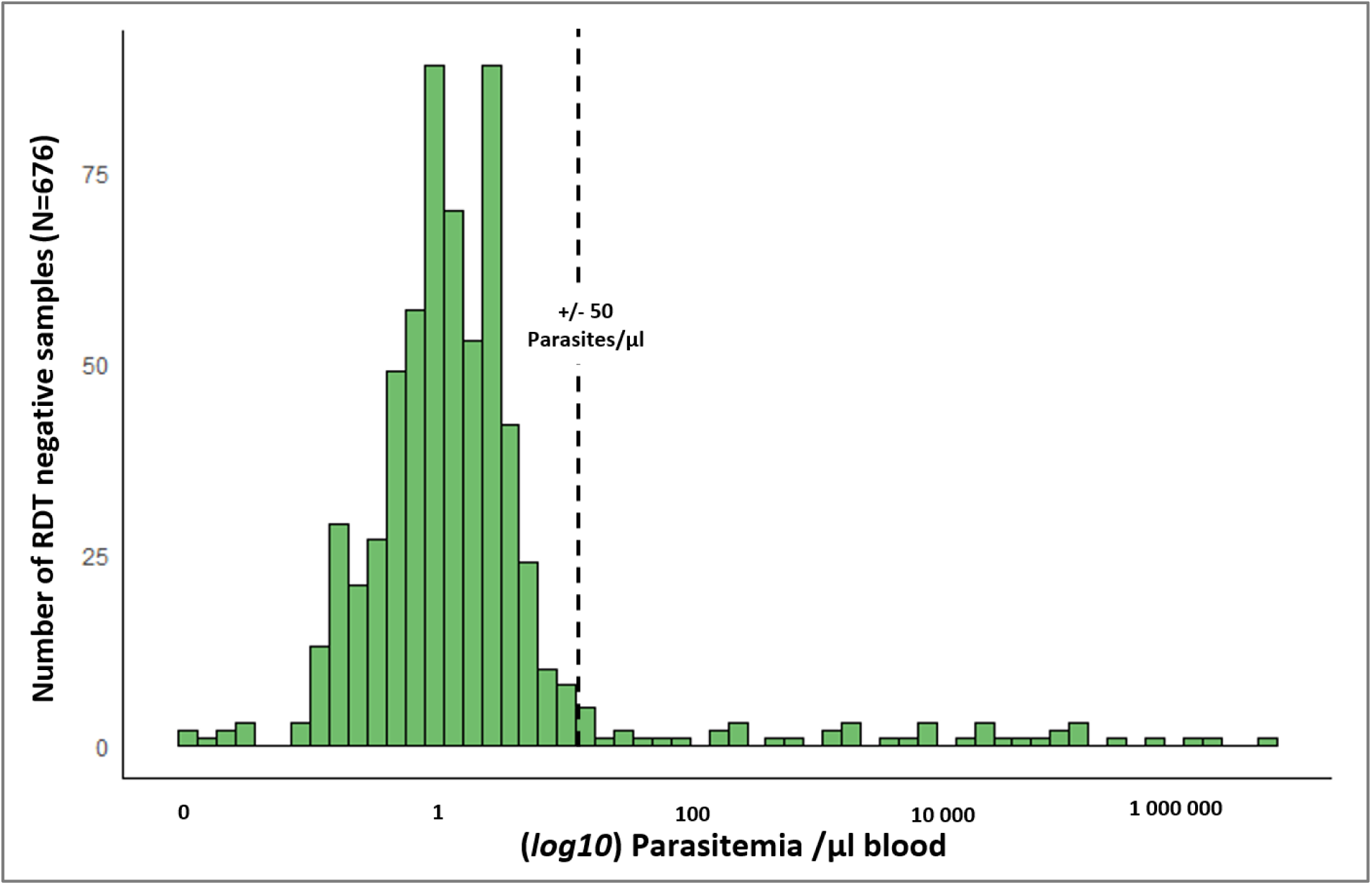
Parasitemia of RDT-negative but qPCR-positive samples detected over the study period

**Figure S3:**
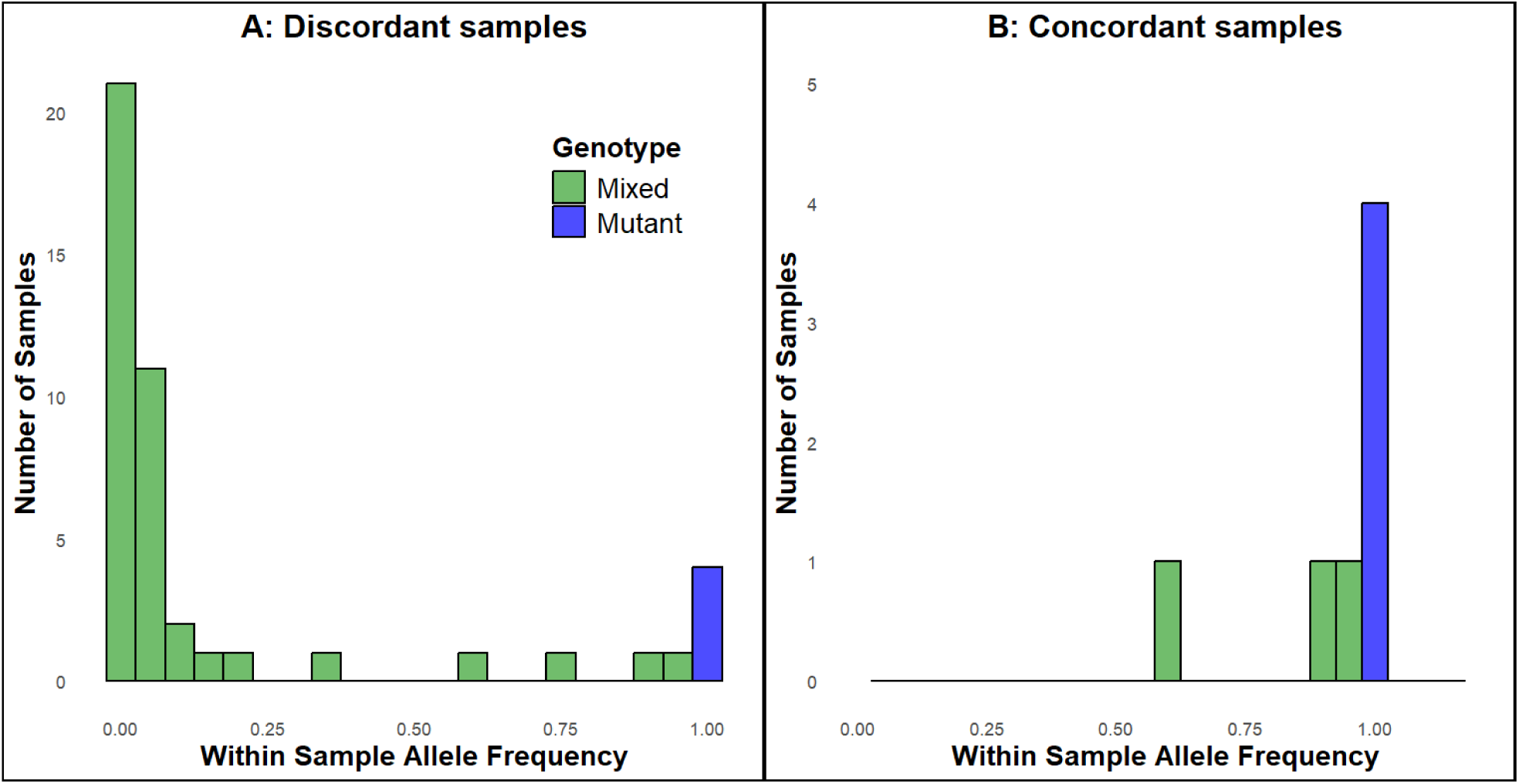
Within Sample Allele Frequencies (WSAF) for A) discordant and B) concordant samples containing a non-synonymous *kelch13* mutation

